# Environmental Surveillance of Bacteria in a New Intensive Care Unit using Plate Sweeps

**DOI:** 10.1101/2025.09.02.25334963

**Authors:** Aasha McMurray-Jones, Kirsten Spann, Prasad Yarlagadda, Jeremy Fernando, Leah W. Roberts

## Abstract

**Background:** The hospital environment plays a critical role in the transmission of infectious diseases. Surveillance methods often rely on selective enrichment or deep metagenomic sequencing, which both have significant drawbacks in terms of community resolution and cost. Plate sweeps provide a practical moderate approach to cultivate a wide range of bacteria, capturing more diversity than a single colony pick without high sequencing costs. Here we use this approach to characterise a newly built hospital intensive care unit (ICU) in Queensland, Australia.

**Methods:** Between November 2023 to February 2024, we sampled 78 sites within a 8-bed private hospital ICU pre- and post-patient introduction to the environment. Samples were enriched on non-selective media before DNA was extracted from whole plate sweeps and sequenced using Illumina. We assessed species, antimicrobial resistance (AMR) genes, virulence genes and transmission across all samples and between the pre- and post-patient samples using Kraken2, AbritAMR, and Tracs.

**Results:** While the rate of positive microbial growth within the ICU environment did not change significantly pre- and post-patient introduction, the post-patient microbiome consisted of largely different bacterial species; of 22 genera identified, only three genera were represented at both timepoints. Post-patient samples were enriched in AMR genes, including resistance to fosfomycin, quinolones, and beta-lactams. Common genera identified post-patient were *Pseudomonas*, *Delftia*, and *Stenotrophomonas*, often associated with areas of plumbing. Cluster analysis identified 17 possible transmission links from a single timepoint, highlighting several areas in the ICU (e.g. communal bathrooms) as key areas for transmission.

**Conclusions:** We demonstrate the utility of plate sweeps as a means of economical non-selective environmental surveillance and highlight its ability to identify hotspots of transmission within a hospital ward that could be targeted by infection control prior to an outbreak of a more serious pathogen.

## Background

Healthcare associated infections (HAIs) are a continuing burden in hospital settings. Annually between 2010 to 2016, ∼83,000 patients were diagnosed with HAIs in Australia^1^, increasing to ∼170,000 in 2018^2^, demonstrating the growing impact of HAIs on Australian patients. Compared to other complications resulting from hospitalisation, infections represent the greatest burden, accounting for 38% of hospital-acquired complications in 2023-2024^3^.

The environment is a key element in seeding and progressing infectious outbreaks in healthcare settings, particularly high-touch (e.g. door handles, keyboards) and plumbing related areas (e.g. bathrooms, sinks)^4,5^. The rise of antimicrobial resistance (AMR) only exacerbates the threat of HAIs, straining our health resources as a consequence of prolonged inpatient stays^6^, intensive infection control requirements, and extra treatment costs^7,8^.

Current environmental surveillance in hospital settings is largely performed phenotypically from single colony picks on media enriched for AMR (Figure 1). This has several limitations, including (i) culture-bias, where only organisms cultivatable in a laboratory setting are identifiable, (ii) resistance-bias, which limits our ability to understand resistance emergence over time, and (iii) limited resolution to track the larger diversity of microbes present at a sampling site. More recently, healthcare settings have utilised metagenomic sequencing directly from environmental samples to capture a complete picture of the microbial community^9^. However, adoption of this method for routine surveillance has been slow, due to prohibitive costs, the complexity of analysing the data, and low-biomass sampling from the hospital environment^9^.

**Figure 1:**
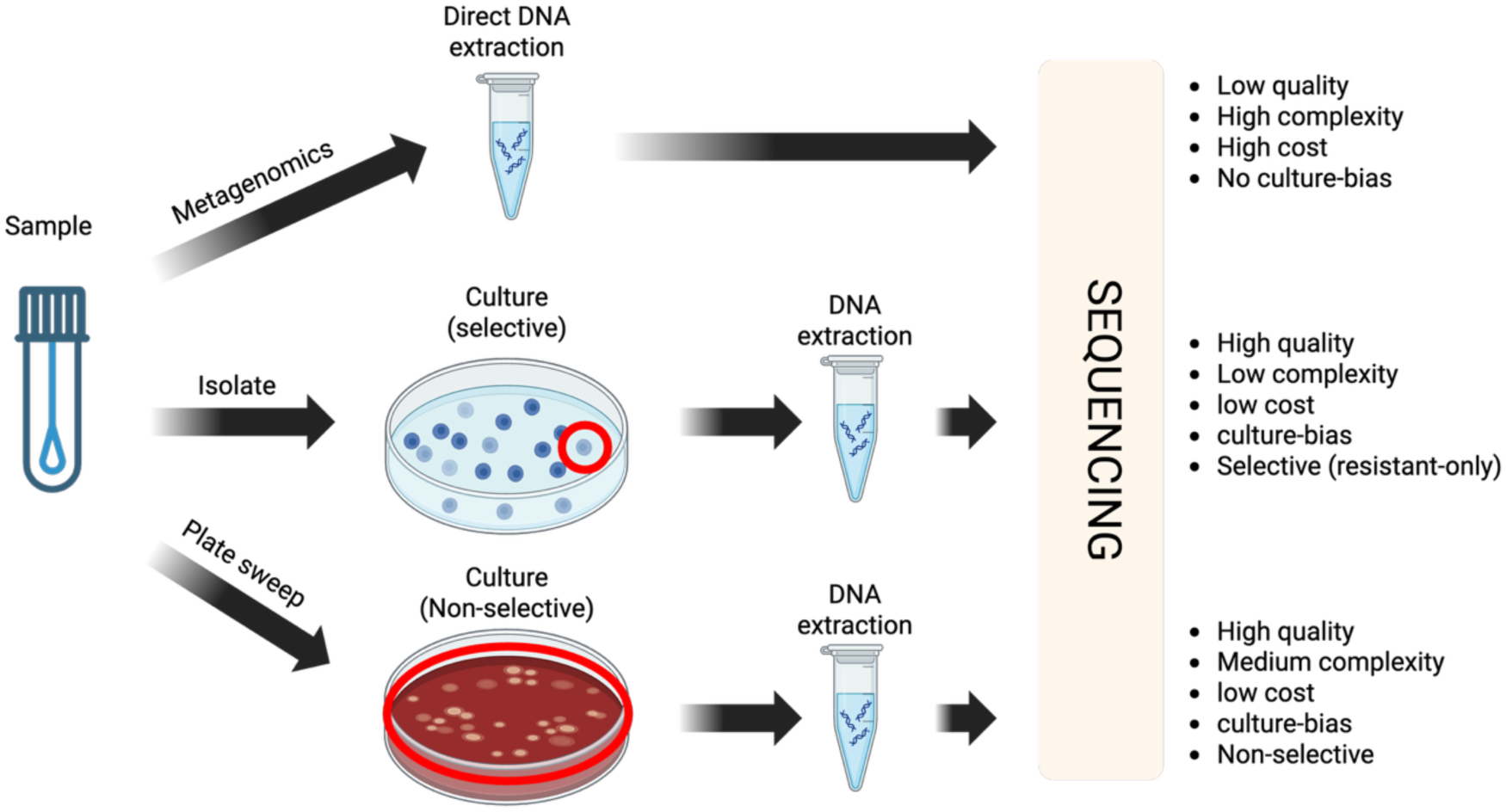
Advantages and disadvantages of environmental surveillance techniques in hospital settings.

As an alternative to these methods, plate sweeps (also referred to as “quasi-metagenomics”) have emerged as an accessible and cost-effective intermediate for understanding bacterial diversity from an environmental sample^10,11^. While plate sweeps are unable to provide a comprehensive community profile (i.e. they still retain culture-bias), most pathogens of clinical interest in the environment are readily culturable^12,13^. Non-selective growth conditions can enrich for a range of bacteria, and DNA extracted from the plate requires less sequencing depth compared to metagenomics. To our knowledge, only one other study has utilised plate sweep techniques for WGS to characterise the hospital microbiome through untargeted surveillance^14^. This approach could provide an important practical step in routine hospital surveillance. Here we conducted a pilot study of the hospital environment using plate sweeps to understand the diversity of environmental bacteria pre- and post-patient introduction.

## Method

### Hospital setting

St Vincent’s Private Hospital, Toowoomba is a regional private hospital with approximately two hundred hospital beds. The new developed intensive care has eight beds including two negative pressure rooms. The main case mix of the ICU includes respiratory failure, sepsis, post-operative care for orthopaedic, ears nose and throat and general surgery. The unit admits approximately 500 patients per year.

### Sampling and storage

Seventy-eight samples from different sites within the new ICU were collected once before patient use (23.11.23 – timepoint 1; T1), and once after patient/staff introduction (22.01.24 – timepoint 2; T2) using standard cotton sampling swabs moistened with sterile water. This included samples from high touch areas (doorknobs, bed frames, sinks, toilets, and medical equipment), areas expected to have high biomass (plumbing and drains), and areas that are infrequently/hard to clean, including keyboards used by staff. Samples were stored in 1 mL of Luria-Bertani (LB) broth at 4℃ for up to 48 hours.

### Culturing, DNA extraction and DNA sequencing

50 μl of swab-LB broth suspension was aliquoted onto each of three agar plates (nutrient (NA), MacConkey (MAC), and Horse Blood agars (HBA)) and incubated at 37℃ overnight. Next, 2 mL of LB broth was added to plates and cultures were resuspended using plate spreaders. 500 μl of resuspended plate sweep was used to extract DNA using DNeasy PowerSoil Pro Kit (QIAGEN) as per manufacturer’s instructions. DNA was sequenced using an Illumina NovaSeq 6000 at the Centre for Microbiome Research, QUT.

### Bioinformatic analysis

Raw sequencing reads were checked for quality using FastQC^15^ (v0.12.1) and trimmed using fastp^16^ (v0.23.4) with “--average_qual 20 --length_required 80” parameters. Human reads were removed using nohuman^17^ (v0.1.1) at default settings. Seqkit^18^ (v2.8.2) was used to assess read metrics pre- and post-filtering.

Trimmed reads were *de novo* assembled using MetaSPAdes^19^ (v3.15.5) and binned using Metabat2^20^ (v2:2.15), at default settings. Species were identified from trimmed reads and binned metagenome assembled genomes (MAGs) using Kraken2^21^ (v2.1.3) and the Kraken2 standard database (20240112). Completeness and contamination for each binned MAG was estimated using CheckM2^22^ (v1.0.1). AMR and virulence genes were identified using AbritAMR^23^ (v1.0.17). Clusters were identified using Skani^24^ (v0.2.1) at a pairwise average nucleotide identity (ANI) threshold >= 99.95 for binned MAGs.

Tracs^25^ (v1.0.1) was used as an end-to-end pipeline for estimating transmission from metagenomic samples. Trimmed and human decontaminated reads from timepoint 2 only were aligned to GTDB (gtdb-rs214-reps.k51.sbt.zip) using the “--keep-all” flag. Two samples (ICU_S39 and ICU_S49) failed as there were zero kmers which aligned to GTDB^26^. Aligned samples were then combined into multiple sequence alignments according to their respective reference genome and distance estimation was performed using the parameters “--snp_threshold 1000 --filter”. Clusters were then inferred with the parameters “--distance filter --threshold 10”.

### Ethics

Low-risk ethics was obtained from St Vincent’s Health and Aged Care Human Research Ethics Committee (HREC_23-18_JFER).

## Results

### Plumbing remains a primary source of bacteria in the hospital environment

Seventy-eight sites around the ICU were sampled, once for timepoint 1 (pre-patient introduction T1) and again for timepoint 2 (post-patient introduction T2). Positivity was measured by observing bacterial growth on any media after overnight culture at 37℃. Several media choices were selected to determine the most appropriate non-selective media for routine surveillance. The amount of growth on NA and HBA was comparable at each site and timepoint, with higher species diversity on the HBA plates (supplementary dataset 1).

Overall, 27/78 sites (35%) were positive in T1, and 44/78 sites (56%) were positive in T2. Sample locations were broadly split into three categories,

1: Patient room (including beds, room handles, room equipment and room switches),
2: High-touch communal surfaces (including all areas in the reception, the laundry trolley, and the medicine room handle), and
3: Plumbing-related (all sinks, toilets, showers and drains in both communal and private patient rooms, including door handles to those rooms).

Across all three categories, there was an increase in site positivity from T1 to T2 (Figure 2). Plumbing-related sites had both the highest positivity rate at both timepoints, and the highest increase in positive sites from T1 to T2 (Figure 2). Across T1 and T2: 24 sample sites remained negative at both timepoints and were mainly door handles and switches (17/24, 71%; Figure 3). 17 positive sites remained positive and included bathroom facilities (shared 5/17; or in private rooms 3/17), and high-touch communal areas (4/17). 27 negative sites (mainly plumbing related, 15/27; or patient room related 11/27) became positive, while 10 positive sites (mainly handles and rails in patient rooms and communal areas 8/10) became negative (Figure 3, supplementary dataset 1).

**Figure 2:**
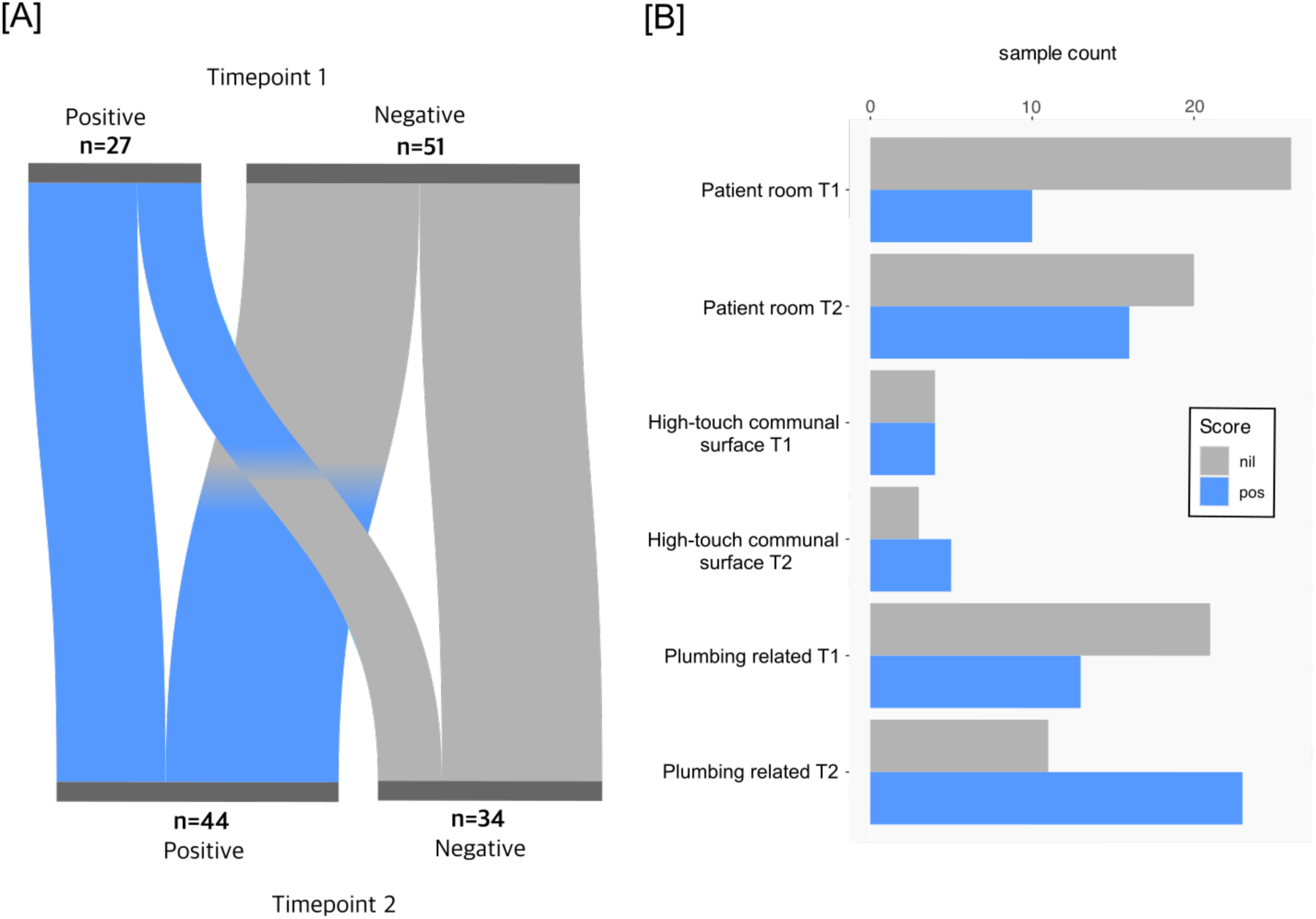
Positive samples by location across Timepoint 1 and 2: (a) a Sankey plot showing change in site positivity/negativity or no change from Timepoint 1 to Timepoint 2. (b) sample counts that were positive or negative for bacterial growth at Timepoint 1 (T1) or Timepoint 2 (T2) by general location (patient room, high-touch, or plumbing). Nil = no growth, pos = positive growth.

**Figure 3:**
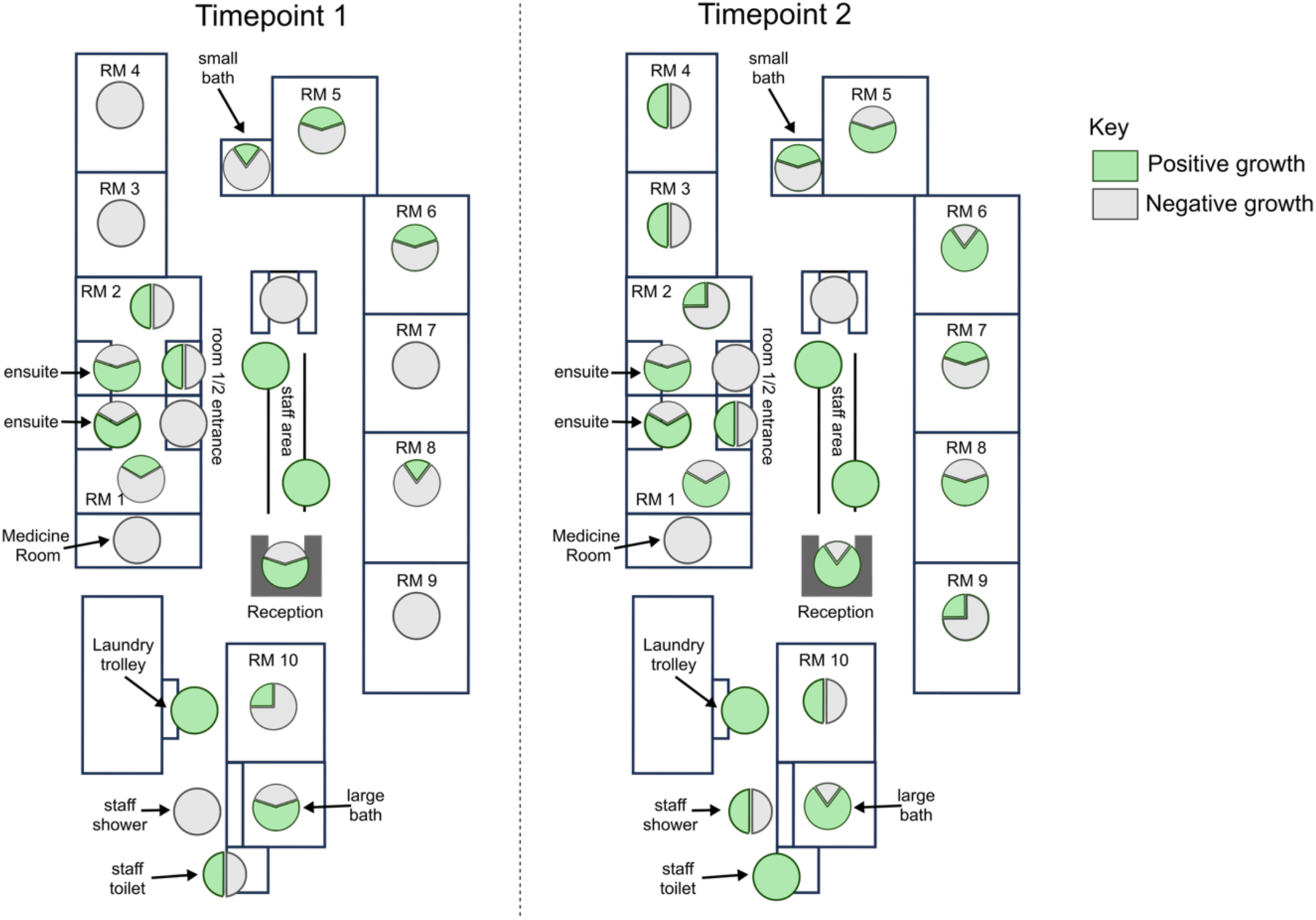
site positivity between T1 and T2: Pie charts showing the proportion of samples from specific regions of the ICU with positive (green) or negative (grey) bacterial growth. Timepoint 1: pre-patient introduction. Timepoint 2: post-patient introduction. RM = patient room. RM 1 and 2 = negative pressure rooms.

### Sequencing revealed abundant changes in the environmental microbiome after patient introduction

Positive samples were triaged for sequencing based on the abundance and type of growth on the media. Of the 27 positive sites for bacterial growth in T1, 15 were sequenced, and of the 44 positive sites for T2, 41 were sequenced (Supplementary dataset 1). To test choice of media, bacteria from all three media types for two samples were sequenced to compare diversity. There was no significant difference in sequences amongst media types, with a slightly higher diversity (although not significantly different) recovered from HBA (supplementary table 1). In addition, filtering to remove human reads had a negligible effect on the total read counts (supplementary dataset 1, supplementary results).

For T1, 13/20 samples could be typed to either the genus or species level, filtering for genus/species assignments above 5%. The exceptions were ICU_S01, which could only be typed to the Bacteria domain, and an additional six samples (ICU_S01, ICU_S03, ICU_S05, ICU_S06, ICU_S08, ICU_S10) that were unable to be classified (>90% unclassified).

Genera identified from T1 were mainly associated with environmental bacteria that have not been routinely reported to cause pathogenesis in humans. This includes *Duffyella, Brevundimonas* and *Cellulomonas* (Figure 4). However, some genera associated with opportunistic infections were also detected, including*, Aerococcus*, *Enterococcus*, and *Pseudomonas*.

**Figure 4:**
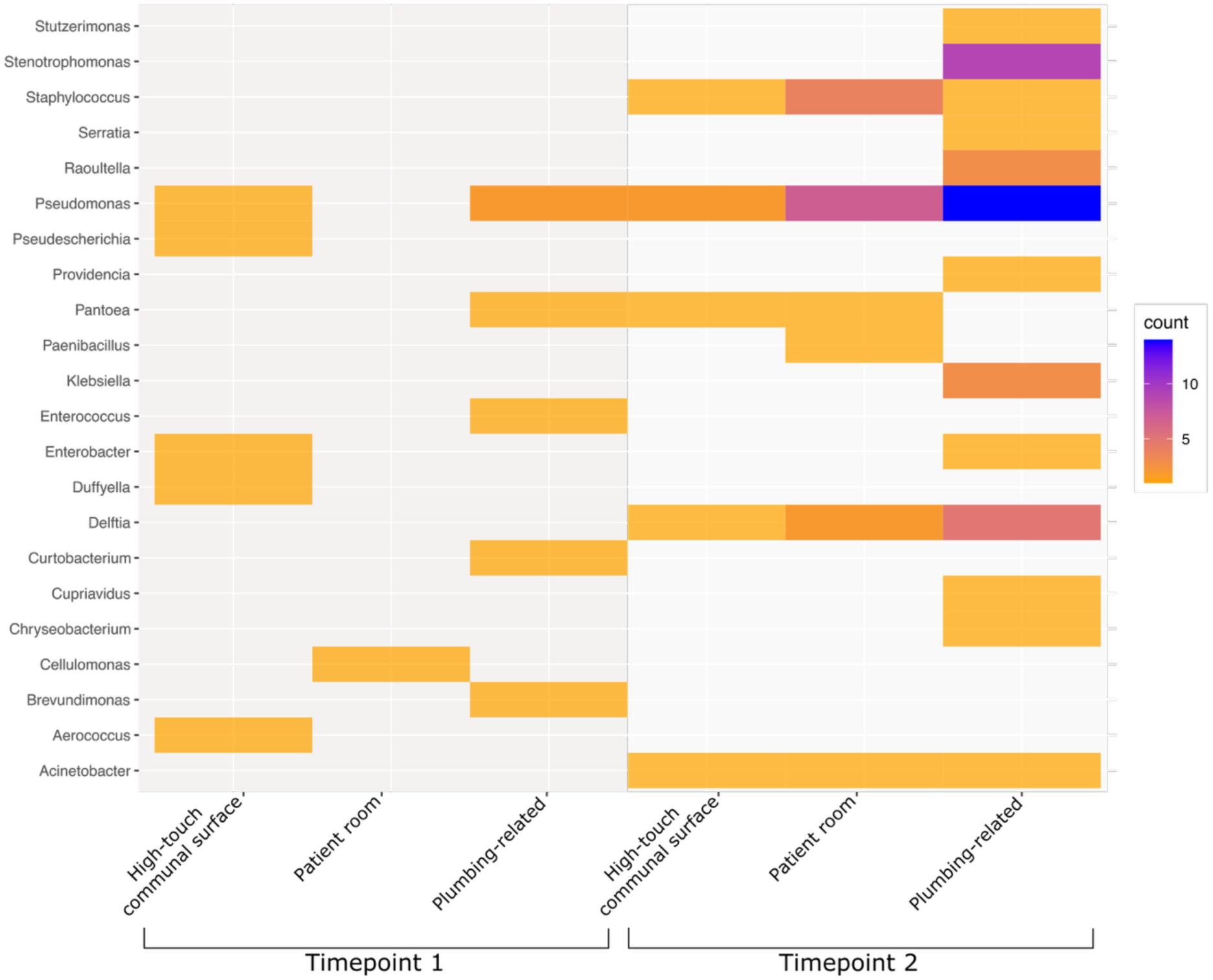
genus identified per location: Sample counts of genera identified for each of the three broad sampling site categories in timepoint 1 and timepoint 2.

**Table 1:**
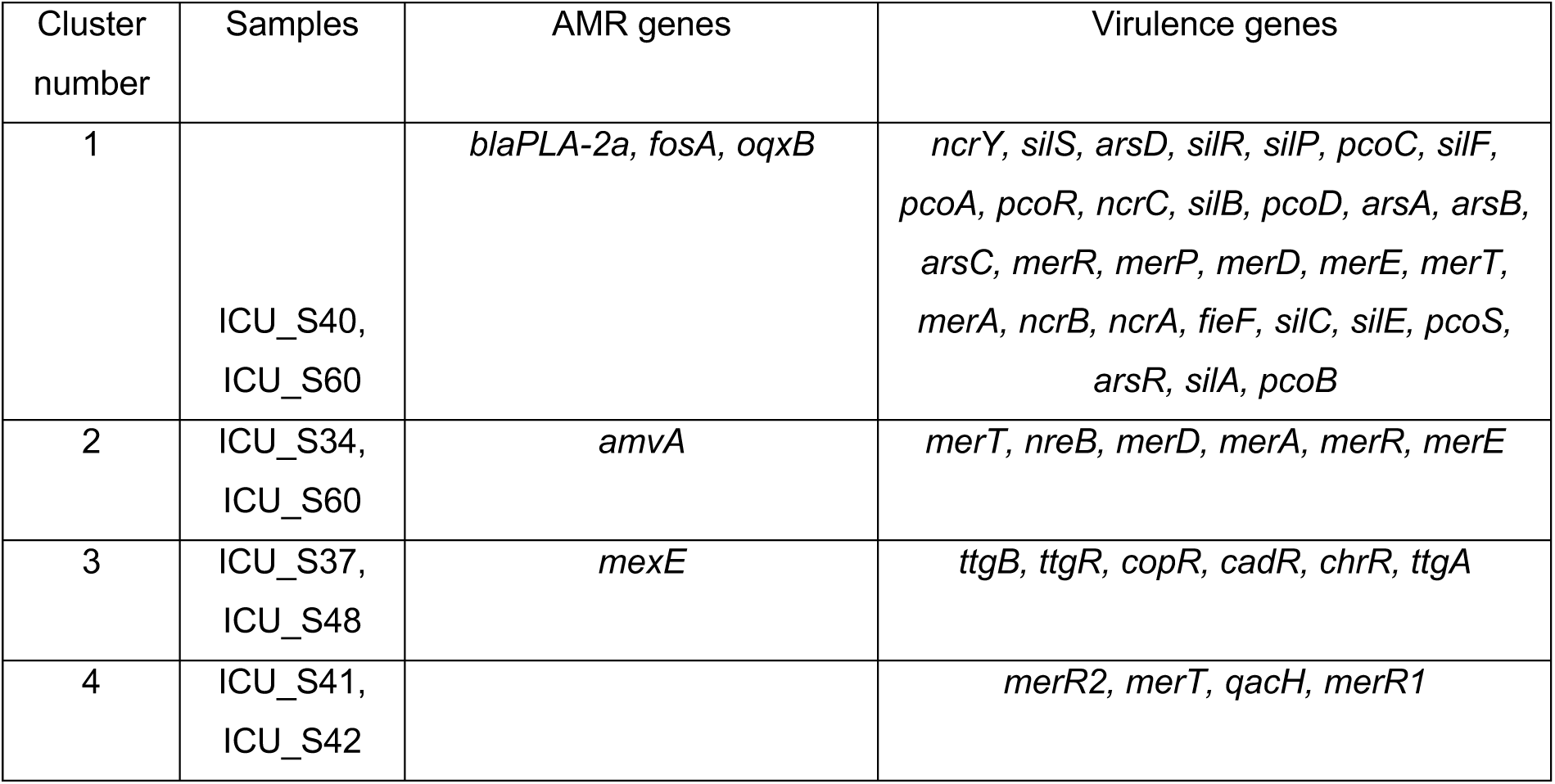
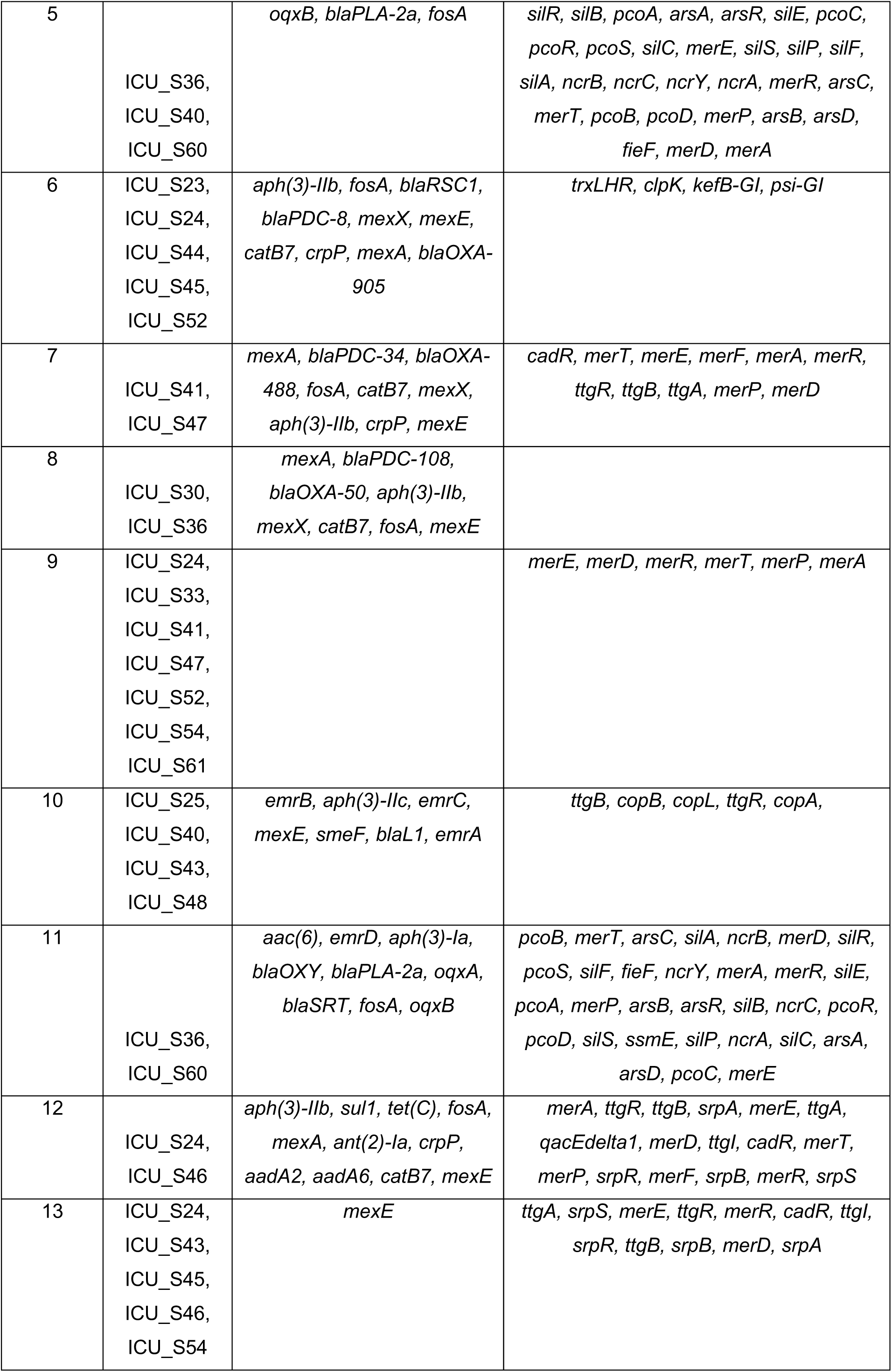
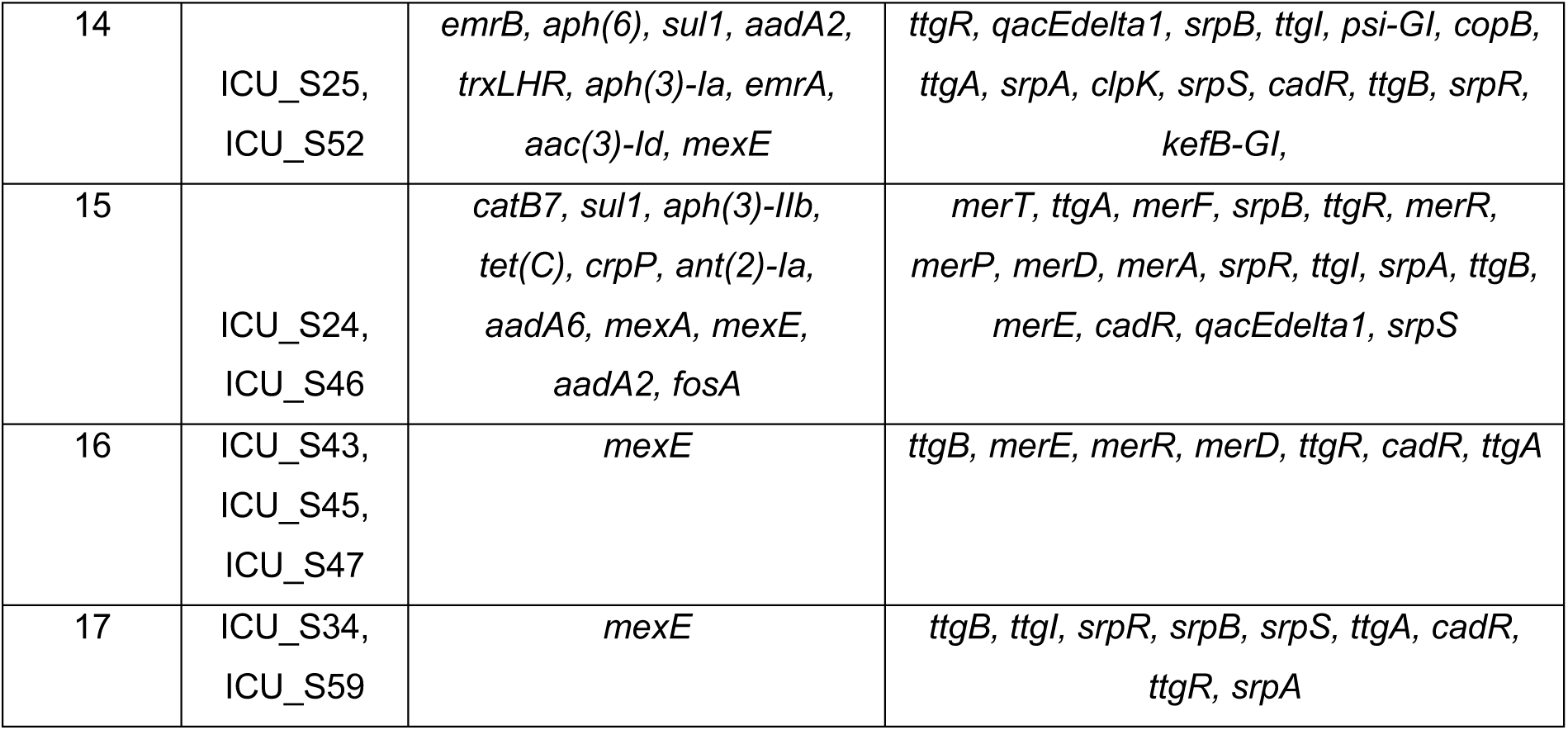
AMR and virulence genes identified across all sampling sites for single clusters.

At T2, two months after patient and staff introduction, we observed a large shift in the microbial environment of the ICU (Figures 4 and 5). Not only were the bacterial residents different, but the rate of positive growth in all areas of the ICU were higher than before patient introduction (Figure 2). Of 41 sites, 39 were positive for at least one bacterial genera at >5% abundance. Two sites (ICU_S39, ICU_S49) were unable to be classified.

**Figure 5:**
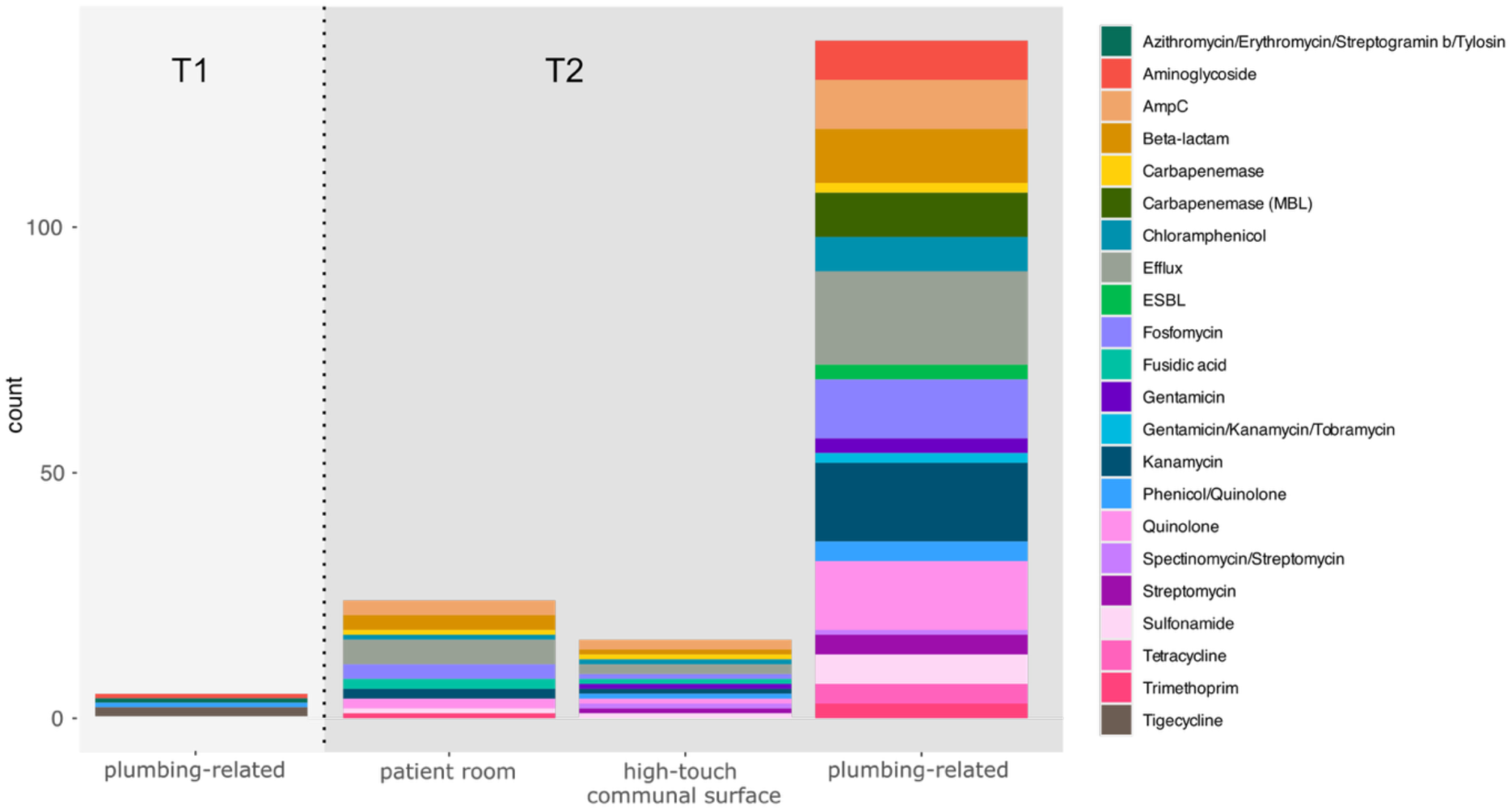
T1 vs. T2 AMR: Number of identified genes across a range of AMR categories detected in T1 and T2 from the three broad sampling site categories.

Twelve additional genera were identified at T2 compared to T1, including well-known human pathogens such as *Acinetobacter, Klebsiella* and *Staphylococcus*. We identified an abundance of *Pseudomonas* (23/41, 56%) and *Stenotrophomonas* (9/41, 22%), which can harbour resistance to last-line antibiotics such as carbapenems, and are frequent residents of hospital environments. Only three genera, *Pseudomonas, Pantoea,* and *Enterobacter,* were represented at both timepoints (Figure 4).

Of four locations where we had paired T1 and T2 sequencing with positive growth (Keyboard 2, Laundry Trolley, Room 1 toilet, room 2 bathroom sink), none had evidence of establishment of the initial T1 microbial community, demonstrating complete displacement by T2. *Pseudomonas* was the dominant genus across all locations, followed by *Stenotrophomonas* and *Delftia* (Figure 4).

Few AMR and virulence genes were identified in T1 samples and only from plumbing-related sites (Figure 5). Two samples (ICU_S07 [room 2 bathroom sink], ICU_S14 [communal bathroom sink]) carried the tigecycline-resistance gene *tmexD2,* a resistance-nodulation-division (RND) family efflux pump. Only two additional samples carried resistance genes; ICU_S15 [room 2 sink] carried *aac(6’)-I* and *msr(C)* conferring resistance to aminoglycoside and azithromycin, and ICU_S16 [room 1 toilet] carried *oqxB9* conferring quinolone resistance. Interestingly, we detected a high prevalence of virulence genes, which consisted mostly of resistance to metals (56% of samples). This included copper (*copB*), mercury (*merABDEFPRT*), silver (*silACPR*) tellurite (*terD*) and divalent metal (*fieF*) resistance. Two genes associated with resistance to cleaning agents, *qacH* and *qacE*, were identified in two samples (ICU_S01 [room 6 bed], ICU_S07 [room 2 bathroom sink]). Overall, at T1 75% of samples (n=12) had no resistance genes detected. 44% (n=7) had no virulence genes nor AMR genes.

Binning of *de novo* assemblies for each sample using Metabat2 enabled closer approximation of the species carrying AMR and virulence genes. Binning of T1 samples identified 111 bins, with 22% (n=24) above 50% completion. *tmexD2* was identified in two *Pseudomonas sp.,* while the *aac(6’)-I* and *msr(C)* genes were identified in an *Enterococcus faecium*, and the *oqxB9* gene identified in *Pantoea agglomerans*.

In contrast to T1, T2 revealed the introduction of substantially more AMR and virulence genes (Figure 5). Efflux pumps were identified in >60% samples, followed by resistance to kanamycin (46%), quinolones (41%), fosfomycin (39%) and beta-lactams (36%). Most samples (80%) carried resistance to metals, followed by biocide resistance (65%). Genes involved in pathogenicity (iron acquisition, biofilm formation) remained rare (n=3). 10 samples (24%) had no AMR genes identified. 5 samples had no virulence genes identified, and 3 samples (ICU_S39 reception bench, ICU_S49 room 5 bed, ICU_S56 room 6 light switch) had neither AMR nor virulence genes.

Once again, binning allowed closer approximation of the species contributing AMR and virulence genes to the sample (Figure 6). In T2 we identified 296 bins, with 43% (n=128) above 50% completion. The efflux pump *mexAEX* was only identified in bins primarily consisting of *Pseudomonas* sp., while *emrABC* was identified in *Stenotrophomonas maltophilia* bins. *S. maltophilia* also appeared to be associated with the carriage of *aph(3’)-IIb* and *smeF*, conferring kanamycin and quinolone resistance. Conversely, *fosA*, *catB7* and *crpP* carriage was more associated with *P. aeruginosa*, although some *Klebsiella* species (*K. pneumoniae, K. michiganensis*) and *Raoltella planticola* also appeared to carry *fosA* along with the efflux *oqxAB*.

**Figure 6:**
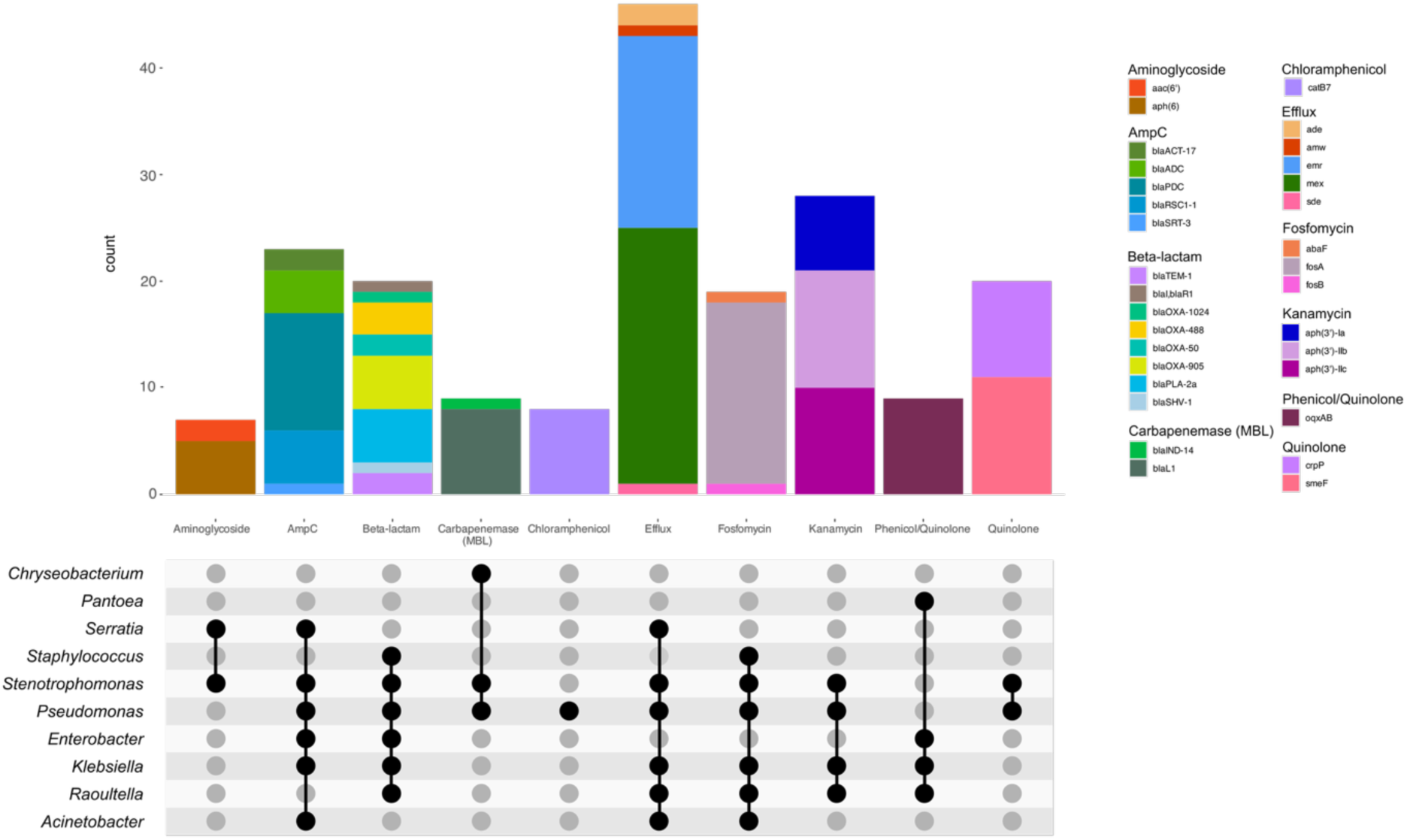
T2 AMR detailed binning: Detailed analysis of AMR genes and predicted associations with genera.

In terms of virulence, *Delftia* sp. was most often associated with mercury resistance (*merADEPRT*), while copper resistance (*copABLR*) was more common in *P. aeruginosa* and *S. maltophilia*. Arsenite efflux (*arsBCR*) was found in a number of species, including *Staphylococcus aureus*, *K. michiganensis*, *K. pneumoniae*, *Escherichia coli*, *Enterobacter hormaechei* and *R. planticola*. Nickel resistance (*ncrABC*) and silver resistance (*SilABEFRS*) was only found in *Klebsiella sp*. with the exception of one sample containing *Pantoea* sp. with *silA*, *silP* and *silR*.

*Pseudomonas* sp. were the main contributors of biocide resistance genes, with *ttgABR* conferring toluene resistance and *sprABRS* involved in gliding motility. Quaternary ammonium compound (qac) resistance genes were identified in a number of species, including *Paenibacillus urinalis*, *Delftia* sp., *Pseudomonas* sp., and *Staphylococcus* sp.

Binning did result in loss of AMR and virulence gene resolution for genes that could not be adequately binned: out of 38 sample sites from T2 that had at least one resistance gene, we found that 22 samples lost AMR or virulence genes through binning, with a median of 5 genes (range 1-30) per sample not reported in binned MAGs.

### Cluster analysis from plate sweeps identifies transmission hotspots

To determine if we could identify transmission from hospital plate sweeps, we utilised two methods for clustering of strains from metagenomic data. The first method relies on assembly and binning of metagenome-assembled genomes (MAGs) followed by rapid average nucleotide identity (ANI) using approximate-mapping with Skani. The second method utilises a mapping and alignment approach using a large reference database to generate strain-specific multiple sequence alignments across samples and then infer single nucleotide polymorphisms (SNPs) using an empirical Bayes approach (Tracs). Given the lack of similarities between T1 and T2, we focused solely on identifying possible transmission of strains across sites in a single timepoint (T2).

To identify high-confidence clusters using Skani, we required MAGs with >70% completeness (based on CheckM), >99% ANI and an alignment length of at least 70%. Using these thresholds, we identified 13 clusters (>=2 samples) from 29 samples (70%). One cluster was removed as we could not identify any known bacterial species in the associated MAGs.

Using a threshold of 10 SNPs with Tracs, we identified 17 clusters from 22 samples (54%). There was clear overlap in the clusters detected between both methods, with 70% of clusters from Tracs and 78% of clusters from Skani identifying the same species/sample combinations (supplementary dataset 1). In particular, *Pseudomonas, Delftia, Paenibacillus,* and *Stenotrophomonas* clusters were identified by both methods. Areas of disagreement were the detection of clusters containing *Acinetobacter* (Tracs only), *Staphylococcus* (Skani only) and *Pseudomonas* (both Skani and Tracs).

Results from the Skani clustering identified possible contamination from the binning step, as some clustered MAGs had several identified species (n=11/51, ∼20%, supplementary dataset 1). As such, we moved forward with interpretation of the Tracs results in the ICU environment (Figure 7). Hotspots for transmission were identified in several locations, notably both shared bathrooms, the negative pressure rooms (room 1 and 2), the staff toilets, the reception desk, and room 8. The two sites linked to the most diverse transmission clusters were ICU_S24 (small bathroom toilet rail) and ICU_S60 (large bathroom sink). The most widespread transmission clusters were cluster 9 (*Delftia tsuruhatensis*, 7 sites), followed by cluster 6 (*Pseudomonas aeruginosa*, 5 sites) and cluster 13 (*Pseudomonas juntendi*, 5 sites).

**Figure 7:**
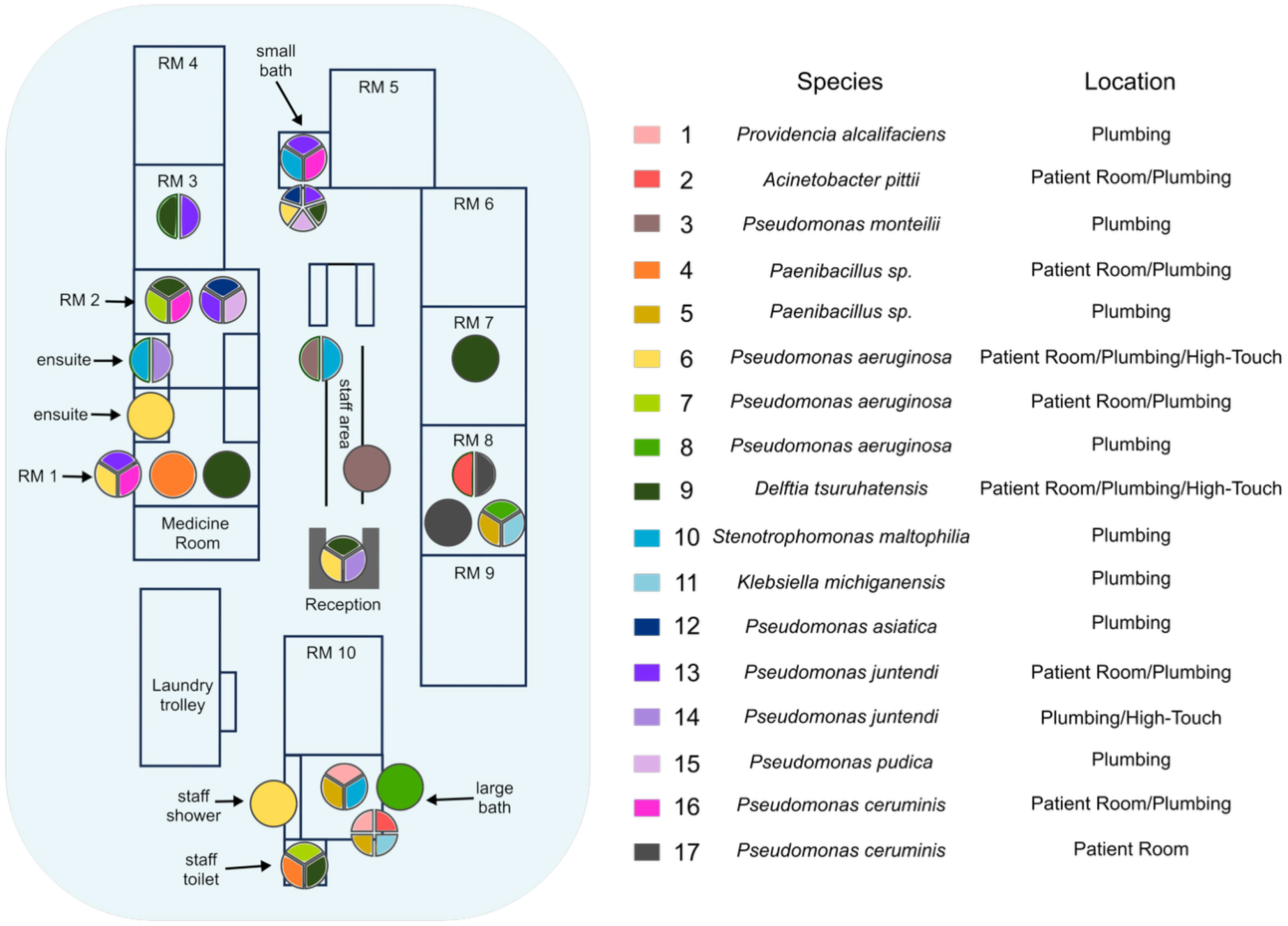
clustering identified using Tracs: Clusters were identified using a single nucleotide polymorphism (SNP) threshold of 10. Pie charts represent specific sites within the ICU with multiple transmission clusters detected.

To identify possible AMR and virulence genes linked to these transmission clusters, we looked at shared AMR genes found across sites with putative transmission. Across 17 clusters, we found on average 5 AMR genes (range 0-11) and 12 virulence genes (range 0-31) shared across sites.

## Discussion

Here we show the rapid change in the environmental microbiome of a newly established ICU pre- and post-patient introduction using a non-selective plate sweep approach. We show that this approach is sensitive enough to detect AMR and virulence genes as well as possible transmission events and could be a powerful tool for proactive surveillance of the hospital environment.

When it comes to understanding the full genetic diversity of a bacterial sample, the limitation of single colony picks is well established. Even for presumed monocultures of a single infectious organism, deep sequencing of plate sweeps has revealed a plethora of genetic variability^27^. Characterising within-species genetic variation has been computationally feasible for more than a decade^28^ and has been used across multiple studies to reveal a more accurate representation of the intra-patient bacterial community. However, in most cases, these approaches have focused solely on understanding genetic variation of single bacterial species. Indeed, tools such as mSweeps^29^ and to some extent mGEMS^30^ lean towards a narrow focus on particular species, usually accomplished via growth on selective media^11,31,32^. Non-selective plate sweep approaches in non-patient settings have been performed previously^14^, and often coined “quasi-metagenomics”, particularly in the foodborne pathogen surveillance field^33,34^. Apart from *Mycobacterium tuberculosis*, the leading bacterial species associated with either human pathogenesis and/or environmental transmission in hospitals are readily culturable in laboratory settings, including many of the WHO priority pathogens. As such, employing direct metagenomics over enrichment methods is likely only revealing a small fraction of additional pathogenic potential in the hospital environment. Furthermore, intermediate culture enrichment significantly reduces the cost and manual handling involved compared to standard direct-from-sample metagenomics^35^, making it a more feasible ongoing method of surveillance. While not completed here, enrichment can also lead to more detailed assessment of the microbial community, as the lower complexity/higher coverage samples are more accessible to long-read sequencing technologies such as Oxford Nanopore (ONT), which has no native amplification step, and could be used to generate complete length reference genomes and mobile elements (such as phage and plasmids)^14^.

In this study of the ICU pre- and post-patient introduction, we highlight two main findings: (1) microbes were present in the environment pre- and post-patient introduction, but the post-patient environment replaced the initial microbial communities, bringing with it substantial AMR capabilities, and (2) regardless of timepoint, virulence capacity, particularly metal resistance, was essential. To date, several studies have observed the change in environmental microbiomes in new or refurbished healthcare settings^36,37^, with a primary focus on methicillin resistant *Staphylococcus aureus*^38,39^ and Enterococci^40^. Here, we show a diverse range of bacteria can be tracked in the hospital environment without selective pressure. While not applied here, an advantage of our method is that it is possible to observe acquisition of resistance genes in pre-existing sensitive environmental microbes, thus noticing evolution of AMR in the hospital environment. Additionally, we show that identifying transmission hotspots using non-resistant microbes can also provide critical information to infection control staff prior to an outbreak of a more serious pathogenic bacterium.

With the recent development and release of Tracs^25^, we show its utility in providing clear and actionable transmission links from hospital environment plate sweep data. We found that binning with Metabat2 was insufficiently sensitive to provide clear transmission information, despite sharing similarities in clustering to Tracs. We considered the use of Semibin2^41^, however the requirement for a trained machine learning model was somewhat impractical for our purposes, as we did not have prior knowledge of all species expected to be encountered in the hospital environment. An additional technical limitation we discovered was that while binning improved estimation of origin for some resistance genes, many resistance genes were also lost due to the inability to confidently place them (and other mobile genetic elements) into appropriate species bins.

Unsurprisingly we found that plumbing was a major source of pathogenic bacteria and AMR/virulence genes. This has been reported numerous times across various healthcare settings^42,43^. We also found a number of key species routinely detected in hospital environments, including *P. aeruginosa*^44^ and *S. maltophilia*^45,46^. The rapid acquisition of these opportunistic pathogens, which contain intrinsic resistance to a number of antimicrobials, may assist the survival of other species in the hospital environment. *Pseudomonas* sp. in particular were found in conjunction with *Acinetobacter*, *Stenotrophomonas*, *Klebsiella*, *Raoultella*, *Serratia* and *Brevundimonas*, and has been implicated in co-infections with other microbes due to its proclivity for biofilm formation^47^.

There were several limitations of this study. Firstly, we were unable to collect samples from patients or continue environmental surveillance beyond our two timepoints. As such, we do not know what the stability of this environmental microbiome will be over time, or how tightly linked it is to the patient cohort. Secondly, we could not perform isolate or metagenomic sequencing alongside to determine the robustness of the plate sweep approach in providing a suitable approximation of the pathogenic potential within the environment. We also acknowledge that competition and overgrowth on the plates will disrupt the actual proportion of bacteria and is therefore not representative of the real population ratios. We did not explore anaerobic growth as a condition but acknowledge that this could be an easily incorporable element to this type of surveillance.

In summary, we present a pilot study demonstrating hospital environmental surveillance using plate sweeps in a newly built ICU in regional Queensland, Australia. We identified rapid colonisation of the ICU environment post-patient introduction with multiple pathogenic bacteria. Non-selective enrichment enabled tracking of bacteria around the ICU, identifying key hotspots for transmission linked mainly to bathrooms. Implementation of this surveillance is cost effective enough to be performed routinely and can identify target areas for additional infection control measures before an outbreak takes place.

## Supporting information

Supplementary Results

## Acknowledgements

We wish to acknowledge the staff at St Vincent’s Private Hospital Toowoomba for accommodating our sampling trips to the intensive care unit. We also acknowledge Michael Hall for advice and readership on the final manuscript.

## Data availability

Reads (post human read removal) have been uploaded to the SRA under Bioproject PRJNA1310799.

## Contributions

Conceptualisation: JF, PY, KS, LWR

Sample collection: LWR

Ethics: LWR, KS, JF

Formal analysis: AMJ, LWR

Funding acquisition: KS, PY, LWR

Investigation: AMJ, LWR

Methodology: AMJ, KS, LWR

Project administration: KS, LWR

Resources: LWR

Supervision: LWR

Visualisation: AMJ, LWR

Writing – original draft: AMJ, LWR

Writing – review & editing: AMJ, LWR, KS, JF, PY

