## Supplementary Results for "Environmental Surveillance of Bacteria in a New Intensive Care Unit using Plate Sweeps"

### Supplementary Data


**Authors:**

Aasha McMurray-Jones[1], Kirsten Spann[1], Prasad Yarlagadda[2], Jeremy Fernando[3], Leah W. Roberts*[1,4,5]

1. Centre for Immunology and Infection Control, School of Biomedical Sciences, Queensland University of Technology, Brisbane, Australia
2. University of Southern Queensland
3. St Vincents Private Hospital, Toowoomba, Queensland, Rural Clinical School, The Medical School, University of Queensland
4. The University of Queensland, UQ Centre for Clinical Research, Herston, QLD 4029, Australia
5. Australian Infectious Disease Research Centre, University of Queensland


#### Supplementary Results:

**Supplementary table 1: species identified after sequencing growth on differential medias**

| | | | |
|---|---|---|---|
| **SML bath sink** | ICU_S12 – NA | ICU_S13 – MAC | ICU_S14 - HBA |
| | *Pseudomonas alcaliphila*<br>*Brevundimonas sp.* | *Pseudomonas alcaliphila*<br>*Pseudomonas oleovorans*<br>*Pseudomonas chengduensis* | *Pseudomonas sp.*<br>*Brevundimonas pondensis* |
| **Keyboard 2** | ICU_S18 – NA | ICU_S19 – MAC | ICU_S20 - HBA |
| | *Duffyella gerundensis* | *Duffyella gerundensis* | *Duffyella gerundensis*<br>*Pseudomonas sp.* |

#### Results of read filtering:

Sequencing of samples produced on average 3.5gb and 3gb of data per sample for T1 and T2, respectively. Filtering for quality removed on average 11% (T1) and 14% (T2) of overall bases. Human decontamination had a negligible effect, removing on average 0.5% and 0.1% of additional bases from filtered reads for T1 and T2 samples (supplementary dataset 1).
